# Improved diagnosis of SARS-CoV-2 by using Nucleoprotein and Spike protein fragment 2 in quantitative dual ELISA tests

**DOI:** 10.1101/2021.04.07.21255024

**Authors:** Carolina De M. Verissimo, Carol O’Brien, Jesús López Corrales, Amber Dorey, Krystyna Cwiklinski, Richard Lalor, Jack M. Doyle, Stephen Field, Claire Masterson, Eduardo Ribes Martinez, Gerry Hughes, Colm Bergin, Kieran Walshe, Bairbre McNicholas, John G. Laffey, John P. Dalton, Colm Kerr, Sean Doyle

## Abstract

The novel Coronavirus, SARS-CoV-2, is the causative agent of the 2020 worldwide coronavirus pandemic. Antibody testing is useful for diagnosing historic infections of a disease in a population. These tests are also a helpful epidemiological tool for predicting how the virus spreads in a community, relating antibody levels to immunity and for assessing herd immunity. In the present study, SARS-CoV-2 viral proteins were recombinantly produced and used to analyse serum from individuals previously exposed, or not, to SARS-CoV-2. The nucleocapsid (Npro) and Spike subunit 2 (S2Frag) proteins were identified as highly immunogenic, although responses to the former were generally greater. These two proteins were used to develop two quantitative ELISA assays that when used in combination resulted in a highly reliable diagnostic test. Npro and S2Frag-ELISAs could detect at least 10% more true positive COVID-19 cases than the commercially available ARCHITECT test (Abbott). Moreover, our quantitative ELISAs also show that specific antibodies to SARS-CoV-2 proteins tend to wane rapidly even in patients that had developed severe disease. As antibody tests complement COVID-19 diagnosis and determine population-level surveillance during this pandemic, the alternative diagnostic we present in this study could play a role in controlling the spread of the virus.

## INTRODUCTION

SARS-CoV-2 is a newly emerging member of the *Coronaviridae* (CoV) family, responsible for the Coronavirus disease-2019 (COVID-19) pandemic. It was first identified in December 2019 in Wuhan, Hubei province, People’s Republic of China, after several individuals developed severe pneumonia similar to that caused by SARS-CoV, the virus responsible for the 2003 SARS outbreak in Asia [1, 2]. Person-to-person transmission of the virus resulted in rapid spreading of SARS-CoV-2 worldwide. More than one year later, the World Health Organization (WHO) reported that SARS-CoV-2 was responsible for more than 130 million infections and 2.8 million deaths around the world [3].

SARS-CoV-2 is an enveloped virus that contains a single-stranded positive-sense RNA. The virus attaches to pulmonary cells via the angiotensin converting enzyme 2 (ACE-2) receptor mediated by a glycoprotein expressed on its surface, the Spike protein (Spro) [4]. Fusion of the viral membrane with the lumen of the endosomal membrane leads to endocytosis, facilitating infection via entry of the viral RNA into the cytosol. During the intracellular viral life cycle, two large polyproteins, pp1a and pp1ab, are translated. Sixteen non-structural proteins (nsp) are co-translationally and post-translationally released from pp1a and pp1ab upon proteolytic activity of two virus proteases, the papain-like protease (PLpro) and the 3C-like protease. These proteins are responsible for the establishment of the viral replication and transcription complex (RTC) which is crucial for virus replication inside the cells [5].

Individuals infected with SARS-CoV-2 can take from one to 14 days to develop symptoms, which range from mild to severe. Common symptoms associated with infection include fever, dry cough, tiredness, loss of taste or smell, aches and pains and diarrhoea. However, infection in a high proportion of individuals can lead to severe acute respiratory syndrome (SARS) or acute respiratory distress syndrome (ARDS) which usually require intensive care. The most severe cases can lead to death [6, 7].

Acute COVID-19 diagnosis mainly relies on real-time reverse transcription polymerase chain reaction (qRT-PCR) or RT loop-mediated isothermal amplification (RT– LAMP) testing of respiratory secretions [8]. In the context of the recent virus variants, whole genome sequencing can also be performed to determine the sequence of the SARS-CoV-2 virus in a sample [9]. Antigen-Detecting Rapid Diagnostic Tests (Ag-RDTs) were developed and have been successfully applied to detect the presence of viral antigens, typically using samples from the respiratory tract to increase the sensitivity of the test [10]. Computed tomography (CT) scans can also be performed and show bilateral multilobular ground-glass opacities which aid in diagnosis. Part of the strategy to identify those exposed to infection and with an established immune response includes serological tests to detect antibodies to SARS-CoV-2. Furthermore, qRT-PCR and serological testing can be used in combination, which was demonstrated to significantly increase the viral detection rates [8, 11].

In general, it takes several days for individuals to build an immune response to the virus. Antibodies to SARS-CoV-2 antigens are detectable in less than 40% of patients within one week of the onset of symptoms, but rapidly increase in the following days [12, 13]. Longitudinal studies are necessary to characterize the longevity of the antibodies in convalescent individuals and to determine if these confer protective immunity [13, 14], and more specifically, to identify which antigen(s) this immunity is directed towards [15, 16]. This knowledge is critical to assess the epidemiological context of the COVID-19 pandemic and for the differentiation between exposed and non-exposed individuals to define the locality and distribution of infection that can guide pandemic control measures such as social distancing. It is also important for vaccine design and the evaluation of vaccine candidates.

SARS-CoV-2 antibody tests are widely used for surveillance studies to gather information about infectivity of the virus in a population. Existing commercial SARS-CoV-2 antibody tests, including enzyme-linked immunosorbent assays (ELISA), chemiluminescence immunoassays and lateral flow assays, were developed using specific viral antigens, principally the Nucleocapsid protein (Npro) and the Spike protein (Spro). The manufacturers of several commercial tests assert that these tests have sensitivities between 86.3 and 100% and specificities from 97 to 100%. However, recent studies that have evaluated the accuracy of antibody tests for use in seroprevalence surveys have reported reduced sensitivities. For example, Schnurra et al. [17] compared the performance of eight different commercial tests and concluded that at least four of them were slightly less sensitive than specified by the manufacturers. Similarly, evaluations made by Public Health England (PHE) found that one in five people with positive results for SARS-CoV-2 in an antibody test used in UK could be wrongly told that they had the infection [18, 19]. Considering the highlighted problems with sensitivity and the limited data regarding the immune response of those individuals beyond 35 days post-symptom onset, such results need to be carefully interpreted by public health authorities [20].

Timely and accurate diagnosis and identification of an immune response to SARS-CoV-2 infection is the foundation of efforts to provide appropriate treatment and recommend isolation that ultimately can contain the COVID-19 pandemic. Npro and Spro-based tests were observed to react with different sets of sera and, therefore, using a combination of viral antigens to assess the antibody response could represent a strategy to increase the accuracy of identifying true positives [17]. In the present study we demonstrate how the current serological diagnosis of SARS-CoV-2 can be improved by using two highly immunogenic virus proteins, Npro and the S2 subdomain of Spro (S2Frag), in a dual ELISA test to detect specific antibody responses to the virus.

## METHODS

### Selecting viral antigens

In the present study, the full length SARS-CoV-2 Spike protein (Spro, ∼135 kDa) and four different sections, Spike protein fragment 1 (S1frag, 1-686, ∼75 kDa), Spike protein fragment 2 (S2frag, 687-1273, ∼54 kDa), the Spike protein fragment 2 prime region (S2Prime, 816-1273, ∼38 kDa) and the receptor binding domain (RBD, 319-542, ∼29 kDa) were selected for recombinant expression (Fig 1A and B) (see also [15]). The entire Npro sequence (2-1269; 50 kDa) was synthesised for recombinant expression (Fig 1C).

**Figure 1.**
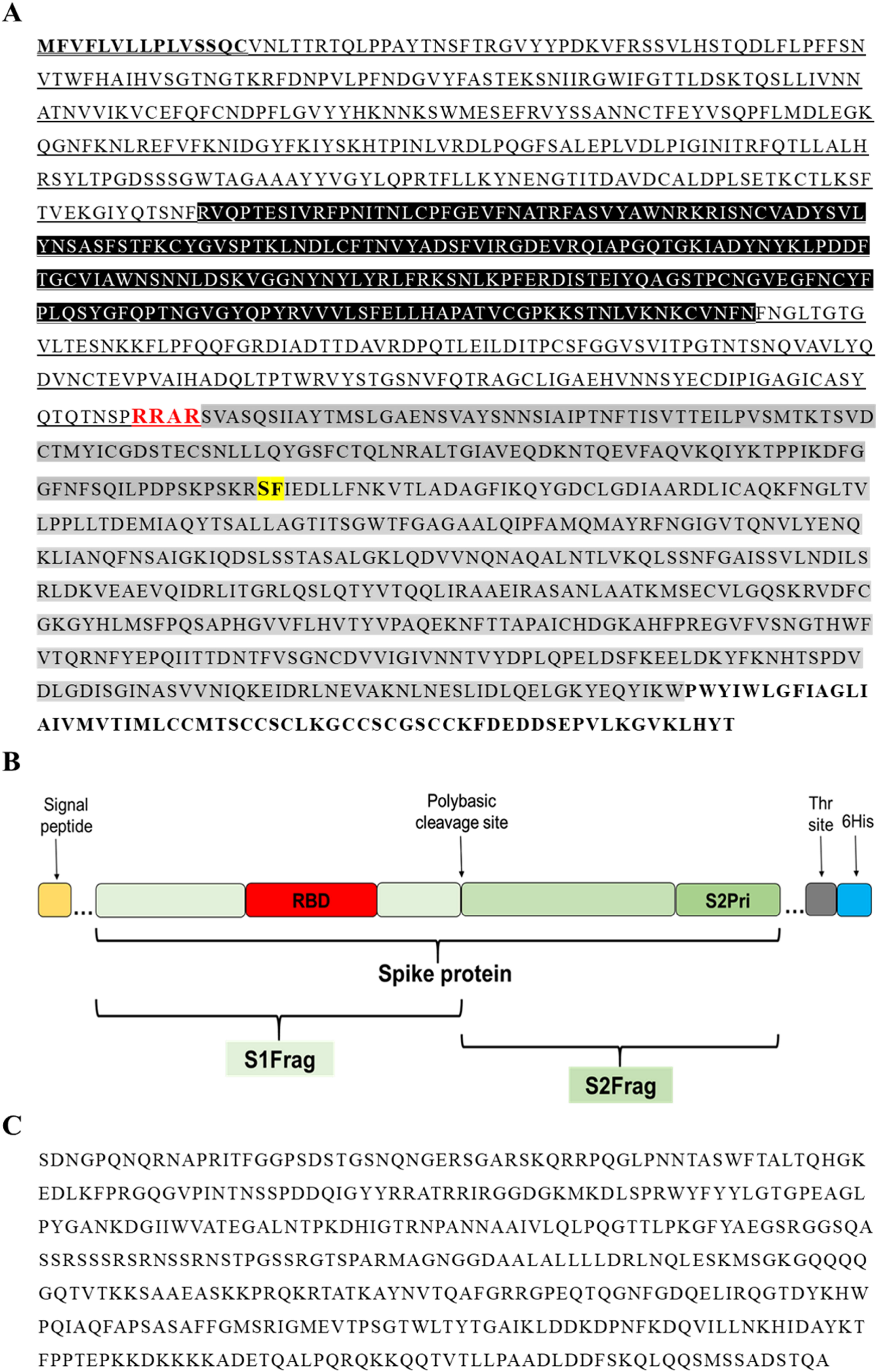
Primary sequence of the SARS-CoV-2 proteins. A: The amino acid sequence of the Spike protein (1273 residues). Residues in bold and underlined represent the signal peptide. Residues highlighted in black (319-542) represent the receptor-binding domain (RBD). Underlined residues delineate the S1-fragment (S1Frag, residue 1-686). Residues in red show the polybasic cleavage site that separates the S1 and S2-fragments (residue 686). Residues highlighted in grey comprise the S2-fragment (S2Frag, residue 687-1273). Residues highlighted in yellow and bold (residue 815) show the beginning of S2Prime sequence (residue 816-1273). Residues in bold represent the transmembrane and endo-domain (1214-1273). B: Schematic representation of the Spike protein and its various portions recombinantly expressed in the present study (see [15]). C: Nucleocapsid protein sequence (Npro, residue 2-1269) used for recombinant expression in *Echerichia coli*.

### Recombinant expression of SARS-CoV-2 proteins in *Escherichia coli*

Sequences encoding the spike protein were codon optimized for expression in *Escherichia coli* and cloned into the pET-28a(+) vector, and into pET-19b for nucleocapsid protein (Genscript Biotech). While Npro contains an N-terminal His-Tag followed by an enterokinase cleavage site, all other proteins contain a thrombin cleavage site followed by a C-terminal His-tag. The synthesized vectors were transformed into BL21 competent *E. coli* cells (ThermoFisher Scientific) following the manufacturer’s instructions and stored in Luria Bertani (LB) broth (Sigma) supplemented with 25% glycerol at −80°C. LB broth supplemented with 50 µg/mL kanamycin, or 100 µg/mL ampicillin for Npro, was inoculated from the glycerol stock and incubated shaking at 37°C overnight. The culture was then diluted in fresh LB broth supplemented with the appropriate antibiotic, incubated at 37°C to OD_600_ 0.6 and protein expression induced with 1 mM isopropyl-β-D-1-thiogalactopyranoside (IPTG; ThermoFisher Scientific) for 4 hr at 30°C. For Npro the cultures were induced with 0.5 mM IPTG for 3 hr at 37°C. Following centrifugation at 10,000 x g for 10 min at 4°C, the bacterial pellets were re-suspended in 10 mL ST buffer (10 mM Tris, 150 mM NaCl, pH 8.0) and stored at −20 °C.

### Solubilisation and purification of recombinant SARS-CoV-2 proteins

Defrosted pellets were treated with 0.1 mg/mL lysozyme in the presence of 40 mM DTT for 1 hr on ice. The proteins in inclusion bodies were solubilised according the protocol described by Schlager et al. [21] protocol. Firstly, a 1% (w/v) SDS buffer (8 mM Na_2_HPO_4_, 286 mM NaCl, 1.4 mM KH_2_PO_4,_ 2.6 mM KCl, 1% (w/v) SDS, pH 7.4) containing 0.1 mM DTT was added to the pellets, which were then sonicated twice for 2 min, 40% amplitude. The samples were centrifuged 15,000 x g at 4°C for 30 min and the resulting supernatant was filtered using 0.45 μm syringe filters. The filtered supernatant containing the soluble recombinant protein was passed through a pre-equilibrated Ni-NTA beads column (Qiagen). The column was washed with 30 mL of wash buffer (8 mM Na_2_HPO_4_, 286 mM NaCl, 1.4 mM KH_2_PO_4,_ 2.6 mM KCl, 0.1% Sarkosyl (w/v), 40 mM imidazole, pH 7.4), and the recombinant protein was eluted using 4 mL of elution buffer (8 mM Na_2_HPO_4_, 286 mM NaCl, 1.4 mM KH_2_PO_4,_ 2.6 mM KCl, 0.1% Sarkosyl (w/v), 250 mM imidazole, pH 7.4). The purified protein was buffer-exchanged into 1x PBS containing 0.05% sarkosyl, pH 7.4.

Recombinant and soluble Npro was extracted from *E. coli* by sonicating twice for 2 min, 20% amplitude (1 g cells: 5 ml lysis buffer (50 mM Tris, 100 mM NaCL, 1 mM EDTA, 10% (v/v) glycerol pH 8.0, with 1 mM PMSF and 4 μg/mL leupeptin), followed by centrifugation and dialysis into 20 mM H_2_NaPO_4_, 500 mM NaCl, 20 mM imidazole pH 7.4. The samples were centrifuged and filtered using 0.45 μm syringe filters, prior to application to HisTrap HP columns (GE Healthcare) equilibrated in the same buffer. After extensive column washing, bound Npro was eluted with 20 mM H_2_NaPO_4_, 500 mM NaCl, 500 mM imidazole pH 7.4. Npro was stored in the elution buffer.

Protein concentrations were verified by Bradford Protein Assay (Bio-Rad) and the proteins visualised on 4-20% SDS-PAGE gels (Bio-Rad) stained with Biosafe Coomassie (Bio-Rad) to check purity. To further confirm the expression and purification of the recombinant proteins, Western blots were performed using a monoclonal mouse anti-polyhistidine antibody (diluted 1:5,000) (Sigma-Aldrich) as a primary antibody followed by incubation with a secondary antibody alkaline phosphatase or horseradish peroxidase (HRP)-conjugated goat to mouse-anti-IgG (diluted 1:5000) (Sigma-Aldrich). Furthermore, the veracity of both S2Frag and Npro recombinant proteins was confirmed by high sensitivity protein mass spectrometry analysis using a Q-Exactive mass spectrometer (ThermoFisher) prior to use for ELISA development [22].

### Human sera samples

Negative controls consisted of a group of 37 serum samples obtained from the Irish Blood Transfusion Service. All the samples were collected prior to SARS-CoV-2 pandemic (2018) and stored at −20°C.

Human serum samples were obtained from St. James’s Hospital, Trinity College Dublin with informed consent. The first set comprised 42 serum samples collected from healthcare workers and all individuals were confirmed to have a SARS-CoV-2 infection by qRT-PCR. All individuals developed symptomatic SARS-CoV-2 infection, and four subjects were hospitalized. The group consisted of 29 females and 13 males, ranging from 27 to 64 years old (average 41.5). The samples were obtained between 17 to 40 days post symptoms onset.

A second set consisted of samples collected from 98 healthcare workers with potential exposure to SARS-CoV-2. This group was divided into symptomatic (N= 49) and asymptomatic (N= 49) individuals. Of the 49 symptomatic individuals, only four were confirmed to have a SARS-CoV-2 infection by qRT-PCR. One of these individuals was hospitalized and admitted to the Intensive Care Unit (ICU). The other 45 individuals were not tested by qRT-PCR because of the number of days after onset of symptoms >7 days. The symptomatic group consisted of 37 female and 12 male individuals, ranging from 23 to 63 years old. The samples were collected between 16 and 113 days after onset of symptoms. The asymptomatic group was formed by 26 females and 23 males, ranging from 22 to 64 years old. Seven individuals in this group were tested doe SARS-CoV-2 infection by qRT-PCR due to close contact status tested and were all given negative results.

Plasma samples from individuals hospitalized with or without COVID-19 related symptoms were obtained. This group consisted of 25 patients, 13 females and 12 males (between 35 to 89 years old), and was divided into qRT-PCR positive (N=15) and qRT-PCR negative (N=10). The plasma samples were collected between 0 and 65 days after onset of symptoms. Two plasma samples, at different time-points, were obtained and analysed from those 15 qRT-PCR positive patients. Of the 15 positive individuals, seven were admitted to the ICU (2 females and 5 males, ranging from 50 to 73 years old). Seven individuals required invasive ventilation. One of the individuals died (male, 79 years old).

Human experimental work was conducted according to Human Research Ethics Committees. Ethical approval for the healthcare worker serum sample collection and analysis was granted by the St. James’s Hospital and Tallaght University Hospital research ethics committee in April 2020 (reference 2020-04 List 15) and permit BSRESC-2020-2403204 (Maynooth University Ethics committee). The work conduced with the samples from hospitalized patients followed the research permit 20-NREC-COV-20 (Galway University hospital research ethics committee). All participants provided written informed consent prior to the study or assent followed by informed consent once able for patients admitted to the ICU where informed consent was not possible.

### Western Blot assays

Purified recombinant proteins (∼2.5 µg/lane) were resolved in a 4-20% SDS-PAGE gel (BioRad) and transferred on to a nitrocellulose membrane. The membranes were incubated in blocking solution (2% BSA-PBST) at 4°C, overnight, then probed with human sera diluted 1:100 in 2% BSA-PBST for 1 hr at room temperature. The membrane was washed four times in PBST before incubation with the secondary antibody, HRP-conjugated goat anti-Human IgG (Fc specific) diluted 1:15000 in 2% BSA-PBST, for 1 hr at RT. The blots were developed for 3 min using 3,3′-Diaminobenzidine substrate (DAB, Sigma-Aldrich).

### Dual antigen SARS-CoV-2 ELISA development

For the dual ELISA tests, separated flat-bottom 96 well microtitre plates (Nunc MaxiSorp, Biolegend) were coated with either Npro (1 µg/mL) or S2Frag (1 µg/mL) diluted in carbonate buffer and incubated overnight at 4°C. The plates were incubated with blocking buffer (2% BSA in PBS-0.05% Tween-20 (v/v), PBST, pH 7.4) and washed. Individual sera samples, diluted 1:100 in blocking buffer, were added in duplicate to antigen-coated wells and incubated for 1 hr at 37°C. After washing five times with PBST, the secondary antibody HRP goat anti-human IgG (Fc specific) (Sigma-Aldrich) was added (1:15,000), and the plates were incubated for 1 hr at 37°C. After washing five times, TMB substrate (3,3′,5,5′-Tetramethylbenzidine Liquid Substrate Supersensitive, Sigma-Aldrich) was added to each well. Following a three-minute incubation the reaction was stopped with 2 N sulphuric acid and plates read at 450 nm in a plate reader (PolarStar). The background value was discounted from the blanks and a cut-off (CO) value for each ELISA test was calculated from the average of all the negative control samples plus three standard deviations. The average OD (450 nm) obtained for each sample tested was divided by the cut-off of the test. Values >1 were considered SARS-CoV-2 positive in the test. Values <1 were negative SARS-CoV-2 in the test. Data were analysed using Prism 5 (Graphpad).

The commercially available Abbott ARCHITECT SARS-CoV-2 IgG ELISA test was performed according manufacturer’s instructions.

### Assessing the antibody response of COVID-19 hospitalized patients using the dual antigen SARS-CoV-2 ELISA

Plasma samples of hospitalized patients with confirmed or suspected COVID-19 infection were tested for the presence of IgG antibodies to Npro and S2Frag using our dual SARS-CoV-2 ELISA assays (see section above). Individual plasma samples, diluted 1:100 in blocking buffer, were added in duplicate into antigen-coated wells and incubated for 1 h at 37°C. The ELISA assays were developed as described above. The average OD (450 nm) obtained for each sample tested was divided by the cut-off calculated for the test. Values > 1 were considered SARS-CoV-2 positive in the test. Values <1 were negative SARS-CoV-2 in the test. Data were analysed using Prism 5 (Graphpad).

### Statistical analyses

Statistical analysis was carried out using GraphPad Prism version 5. Differences between negative controls and positive controls were analysed using an unpaired t-test. Correlation between Npro and S2Frag ELISA tests or Abbott ARCHITECT SARS-CoV-2 IgG ELISA test and our ELISA tests were aanalysed using Spearman’s rank test with 95% confidence intervals.

## RESULTS

### Isolation and solubilisation of SARS-CoV-2 recombinant proteins

Recombinant proteins were successfully expressed in *E. coli* BL21 cells; however, spike protein and its various subunits were associated with insoluble inclusion bodies. By employing a protocol using 1% SDS the inclusion bodies were solubilised and the various proteins purified at ∼1 mg/L of culture (Fig 2). The solubilisation and isolation of S2Frag is shown in Fig 2A and B. Residual insoluble Npro was present in post-lysis recombinant *E. coli* cell pellets. However, high level and soluble Npro expression was observed (yield: 3 mg/L) following affinity chromatography (Fig 2C). High sensitivity proteomic analysis confirmed 53% and 68% sequence coverage for the Npro and S2frag recombinant antigens, respectively (Supplementary Fig S1).

**Figure 2.**
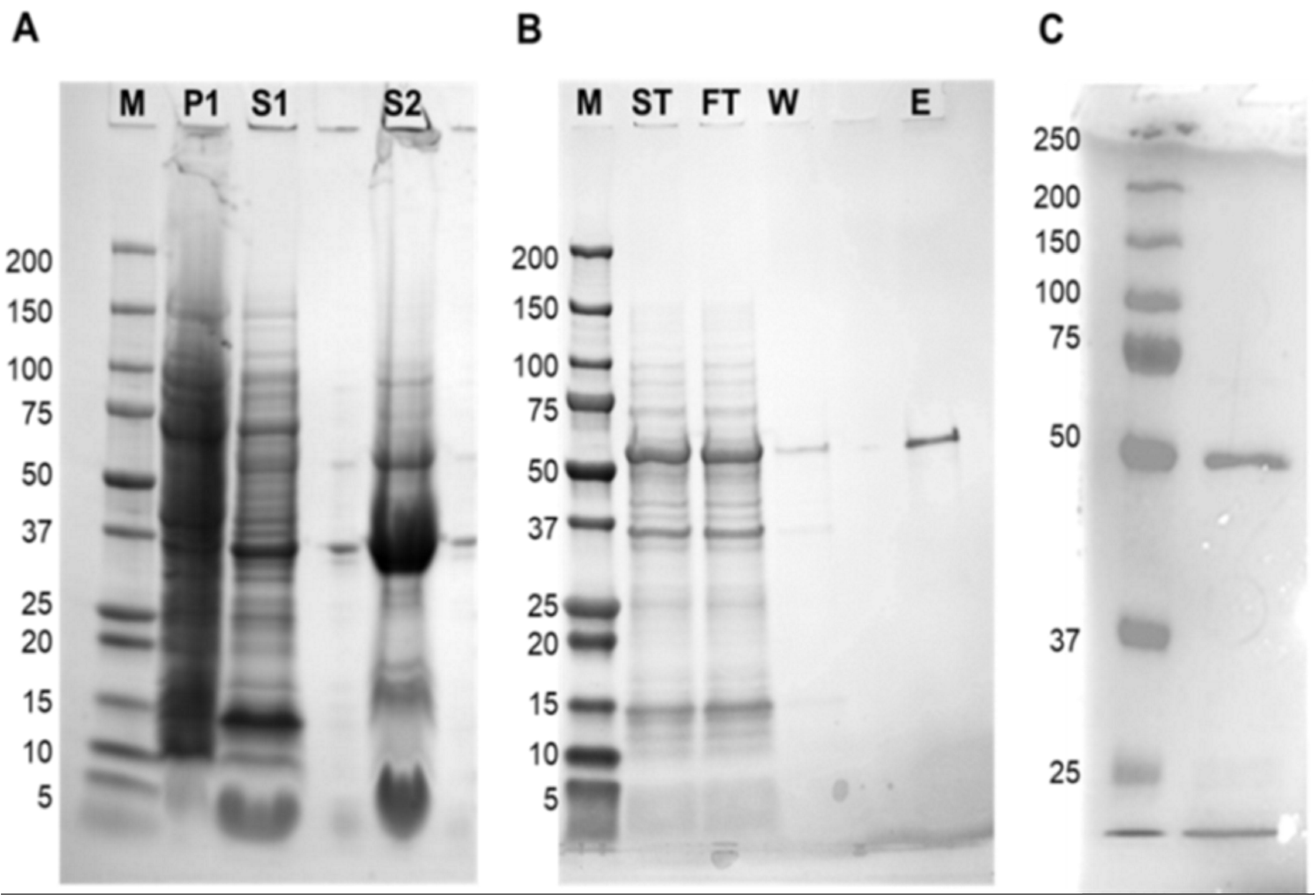
Recombinant production and purification of Spike protein fragment 2 (S2Frag). A: Solubilisation of the S2Frag protein. P1, *E. coli* pellet after induction with IPTG for 4 hr at 30°C; S1, supernatant containing soluble proteins after pellet digestion with 0.1 mg/mL of lysozyme; S2, supernatant containing insoluble proteins after pellet digestion with lysis buffer containing 1% SDS. B: S2Frag purification over Ni-NTA beads column. ST, supernatant total diluted; FT, column flow through; W, washes; E, eluted protein; M, Molecular weight marker in kDa. C. 4-20% SDS-PAGE analysis of recombinant SARS-CoV-2 nucleocapsid protein (Npro) following HisTrap HP columns.

### ELISA antibody test using Npro and S2Frag distinguishes positive and negative SARS-CoV-2 infected individuals

As part of the development and optimization of the in-house ELISA developed with the recombinant Npro and S2Frag, an appropriate cut-off point for each antigen was established using 37 negative control human samples collected pre-COVID-19 pandemic (in 2018). Then, 42 SARS-CoV-2 qRT-PCR positive samples were screened using both Npro-ELISA and S2Frag-ELISA. This screening showed that all 42 individuals assessed generated significant levels of IgG antibodies against both Npro and S2Frag proteins (Fig 3). Notably, infected individuals showed an average antibody response to Npro that was consistently higher than the reactivity against S2Frag. Notwithstanding, both Npro and S2Frag could be employed to distinguish positive and negative SARS-CoV-2 infected individuals.

**Figure 3.**
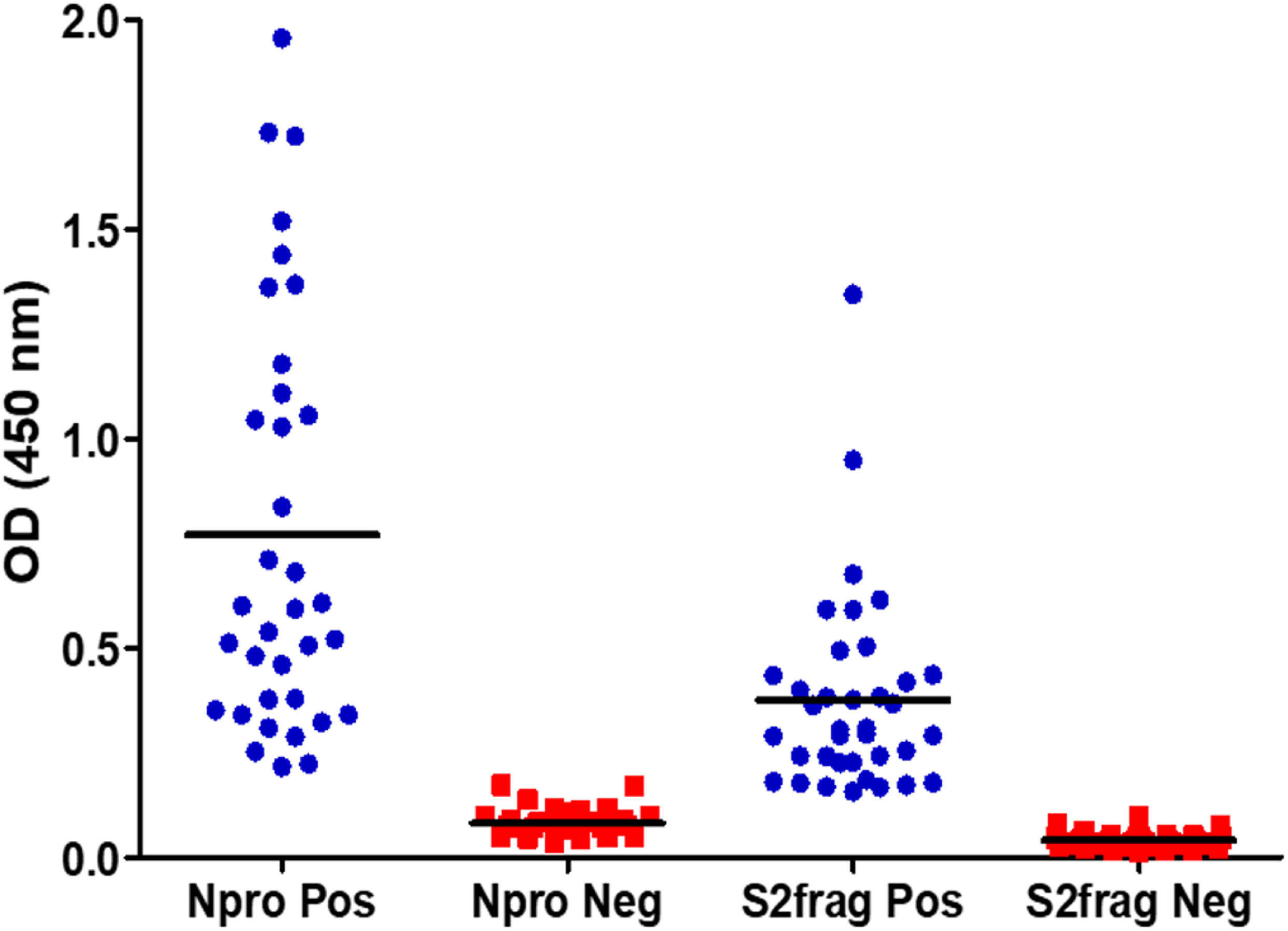
The determination of cut-off values for positive and negative results by ELISA. Forty-two sera samples from patients with a positive SARS-CoV-2 diagnosis by RT-PCR and 37 sera samples stored in a blood bank prior to SARS-CoV-2 were tested by ELISA to determine the cut-off values for a positive or negative result for antibodies against Npro or S2frag. Pos: Positive. Neg: Negative.

Examining the performance of the Npro-ELISA, we deemed 36 (85.7%) samples as positive infected individuals. When these samples were tested with the S2Frag ELISA assay, 37 (88%) positive samples were identified. However, by combining the results of both ELISA tests, the number of positive samples was 40 (95.2%) because not all individuals produced antibodies against both Npro and S2Frag (Fig 4A, Table 1).

**Table 1.**
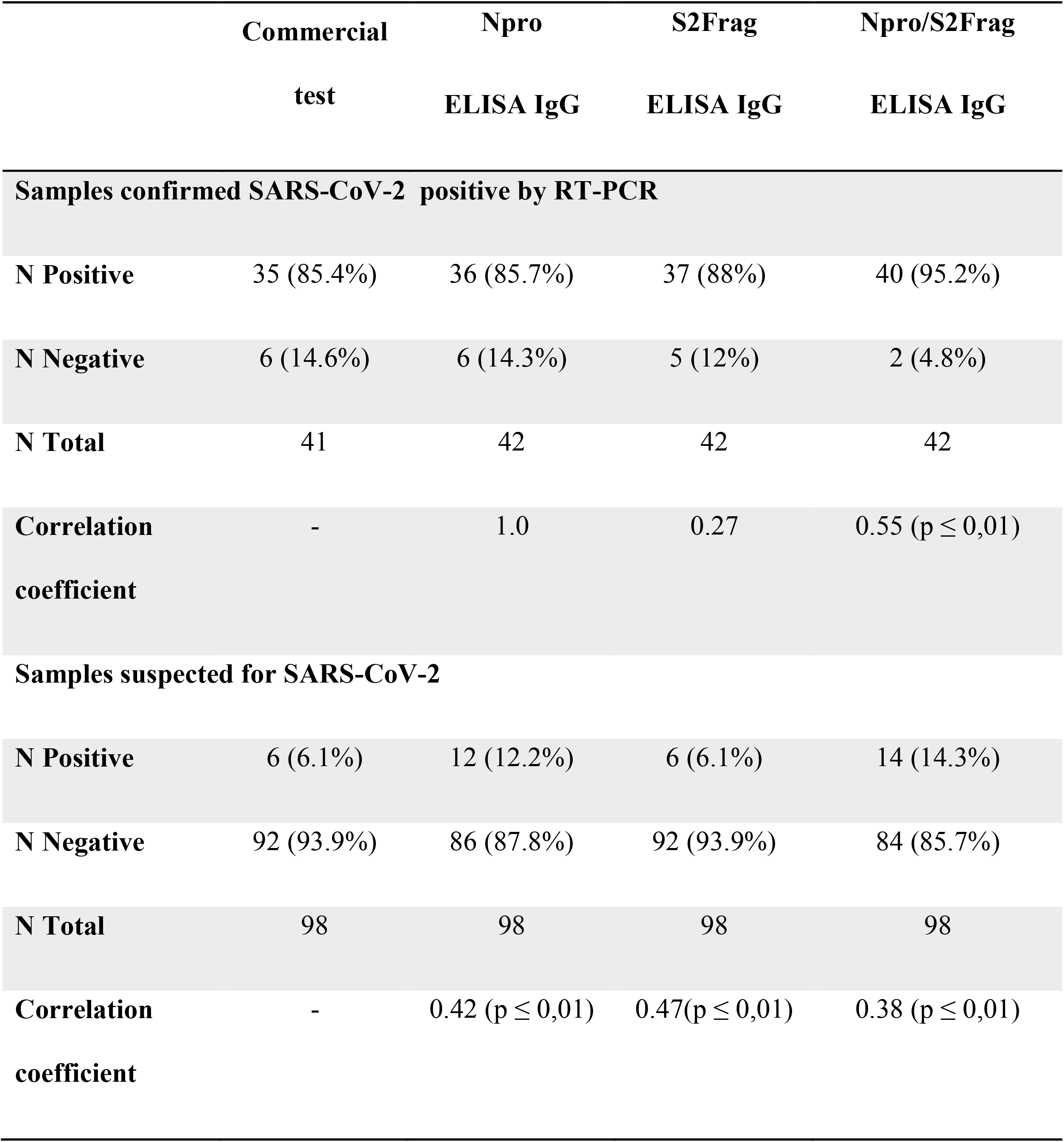
Comparison of the performance of the commercially available Abbott ARCHITECT test and the ELISA developed in the current study.

**Figure 4.**
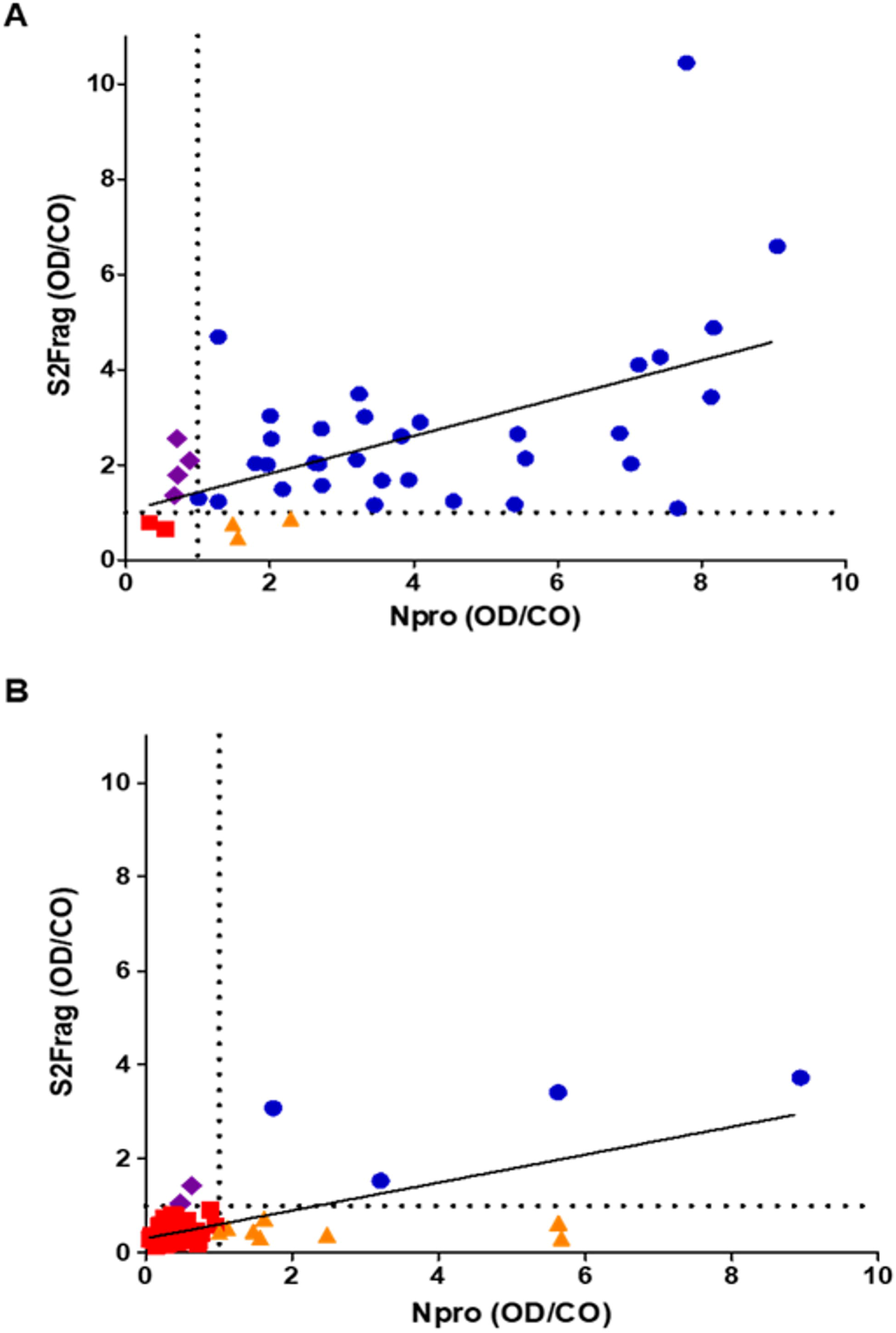
Antibodies against Npro or S2frag detected in sera from individuals confirmed positive for SARS-CoV-2 infection by RT-PCR, or suspected of SARS-CoV-2 infection. (A) Sera from 42 RT-PCR positive SARS-CoV-2 patients were tested for antibodies against Npro and S2Frag by the ELISA antibody test developed in this study. R square: 0.3132. (B) Sera from 98 suspected SARS-CoV-2 individuals were tested for antibodies against Npro and S2Frag. R square: 0.4704. (■ sera were negative for antibodies against both Npro and S2frag by ELISA; ▲ sera were positive for antibodies against Npro only by ELISA; ♦ sera were positive for antibodies against S2frag only by ELISA; ● sera were positive for antibodies against both Npro and S2frag by ELISA). Individual results for Npro and S2Frag ELISA presented as Optical density (OD 450 nm) divided by the calculated cut-off (CO). The cut-off value for each antigen is indicated by the dotted line.

Serum samples from 98 healthcare workers suspected of exposure to SARS-CoV-2 were also screened in both ELISAs. Of these, 12 (12.2%) were detected as positive using only the Npro ELISA (Fig 4B and Table 1) while 14 (14.3%) were deemed positive when the S2Frag ELISA results were considered together with the results of the Npro ELISA.

### Western blot analysis of SARS-CoV-2 positive serum samples

Western blot analysis using purified recombinant Npro and S2Frag proteins was performed on all serum samples. This analysis confirmed infectivity of all individuals that were deemed positive by ELISA. However, a wide range of reactivity was observed between patients, which correlated with our ELISA observations showing that some patients produced antibodies reactive with both Npro and S2Frag while others produced antibodies that reacted with either Npro or S2Frag (see Fig 5 for representative Western blots).

**Figure 5.**
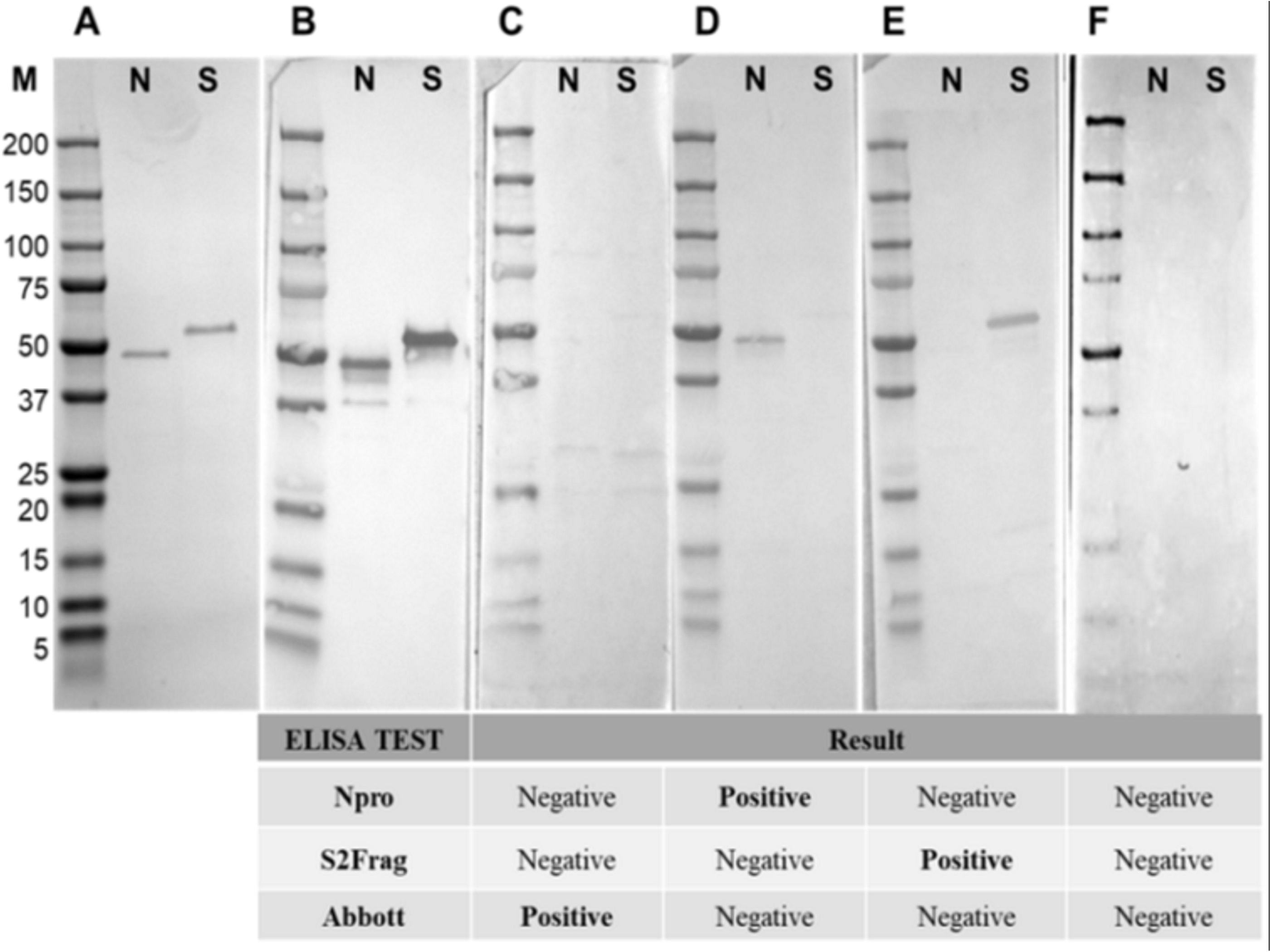
Western blots representative of samples showing the presence and absence of antibodies to Npro and S2frag in individuals positive for SARS-CoV-2. Sera were assayed by Western blot to detect antibodies against Npro (N) and S2frag (S). A: Recombinant proteins resolved in a 4-12% SDS-PAGE and stained with Coomassie-blue. B: Western Blot control performed using a monoclonal mouse anti-polyhistidine antibody (1:10,000) (Sigma-Aldrich) as a primary antibody followed by incubation with a secondary antibody alkaline phosphatase conjugated goat to mouse-anti-IgG diluted 1:5,000 (Sigma-Aldrich). C-F: The antibodies response to Npro and S2frag of different individuals positive for SARS-CoV-2. Individual ELISA tests results are shown for each sample as positive or negative for SARS-CoV-2.

### Comparison of the Npro and S2Frag ELISAs with a commercially available antibody test

In order to assess the sensitivity of the Npro and S2Frag ELISA tests against a commercially available test, serum samples were tested in parallel using the Abbott ARCHITECT ELISA (ARCHITECT SARS-CoV-2) which employs Npro as its antigen (Fig 6). Using the 42 qRT-PCR-confirmed positive serum samples, the data showed complete agreement between the Abbott ARCHITECT and the Npro ELISA test developed in this study (i.e. 85.4% sensitivity) (Fig 6A). However, four patients that were negative and two that were positive by both these tests showed a contrasting result when evaluated by the S2Frag-ELISA (Fig 6B). Combining the data for the Npro-ELISA and S2Frag-ELISA tests increased the sensitivity of detection to 95.2% (Table 1).

**Figure 6.**
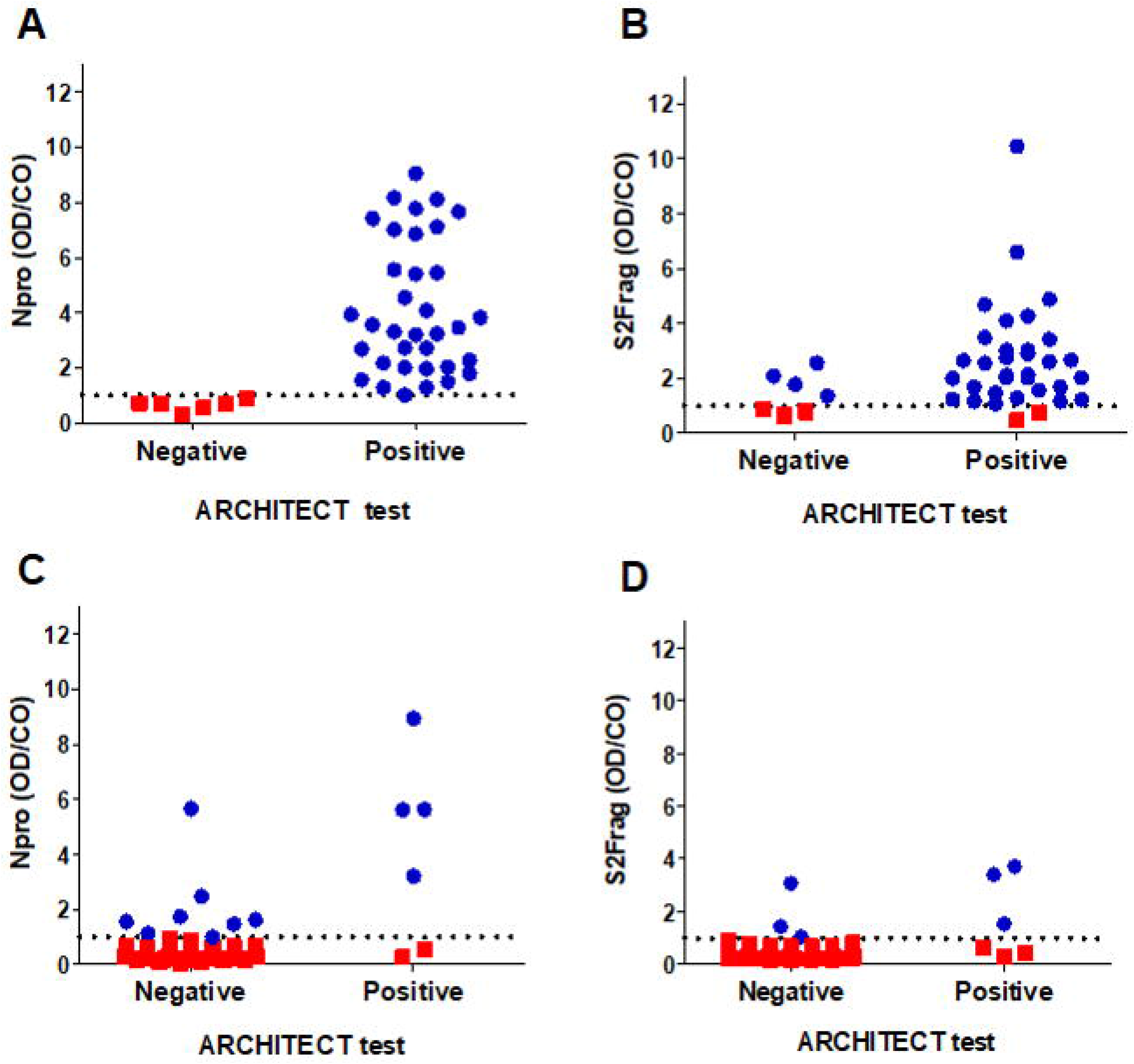
Contrasting results obtained by the commercially available Abbott ARCHITECT antibody test and the ELISA antibody test developed in the current study. A and B, the agreement of SARS-CoV-2 diagnostic of 42 RT-PCR positive SARS-CoV-2 individuals assessed using Npro and S2frag ELISA test or the commercially available Abbott ARCHITECT antibody test. C and D, the agreement of SARS-CoV-2 diagnostic of 98 suspected SARS-CoV-2 individuals assessed using Npro and S2frag ELISA test or the commercially available Abbott ARCHITECT antibody test. Samples were categorised according to the positive or negative result of the commercially available Abbott ARCHITECT test. Individual results for Npro and S2Frag ELISA test presented as optical density (OD 450 nm) divided by the calculated cut-off (CO) (■ sera were negative for antibodies by ELISA; ● sera were positive for antibodies by ELISA). The cut-off value for each antigen is indicated by the dotted line.

When the ARCHITECT test was employed to screen plasma samples from the 98 healthcare workers suspected of exposure to SARS-CoV-2 only six (6.1%) of these samples proved positive for SARS-CoV-2. In contrast, 14 (14.3%) individuals were identified as positive using our in-house ELISA tests (Fig 6C and D and Table 1).

### Antibody responses to SARS-CoV-2 antigens in COVID-19 hospitalized patients

Plasma samples from COVID-19 hospitalized patients were tested for specific antibodies against our in-house Npro and S2Frag-ELISA tests. Two samples of each patient, at different time-points after the onset of symptoms, were assessed and compared for their levels of antibodies against Npro and S2Frag (Fig 7). The data shows that COVID-19 hospitalized individuals develop strong antibody response to both Npro and S2Frag. However, the level of antibody to each antigen is very distinct. The OD/CO values obtained to the Npro were consistently higher than to the S2Frag (Medium OD/CO Npro= 8.46 and S2Frag= 2.09). Moreover, antibodies to Npro could be detected from day seven after onset of symptoms, whilst antibodies to S2Frag were only detected from day 11 (Supplementary Table S1). Nevertheless, from day 15 after onset of symptoms, all individuals assessed showed strong antibody response to both SARS-CoV-2 antigens.

**Figure 7.**
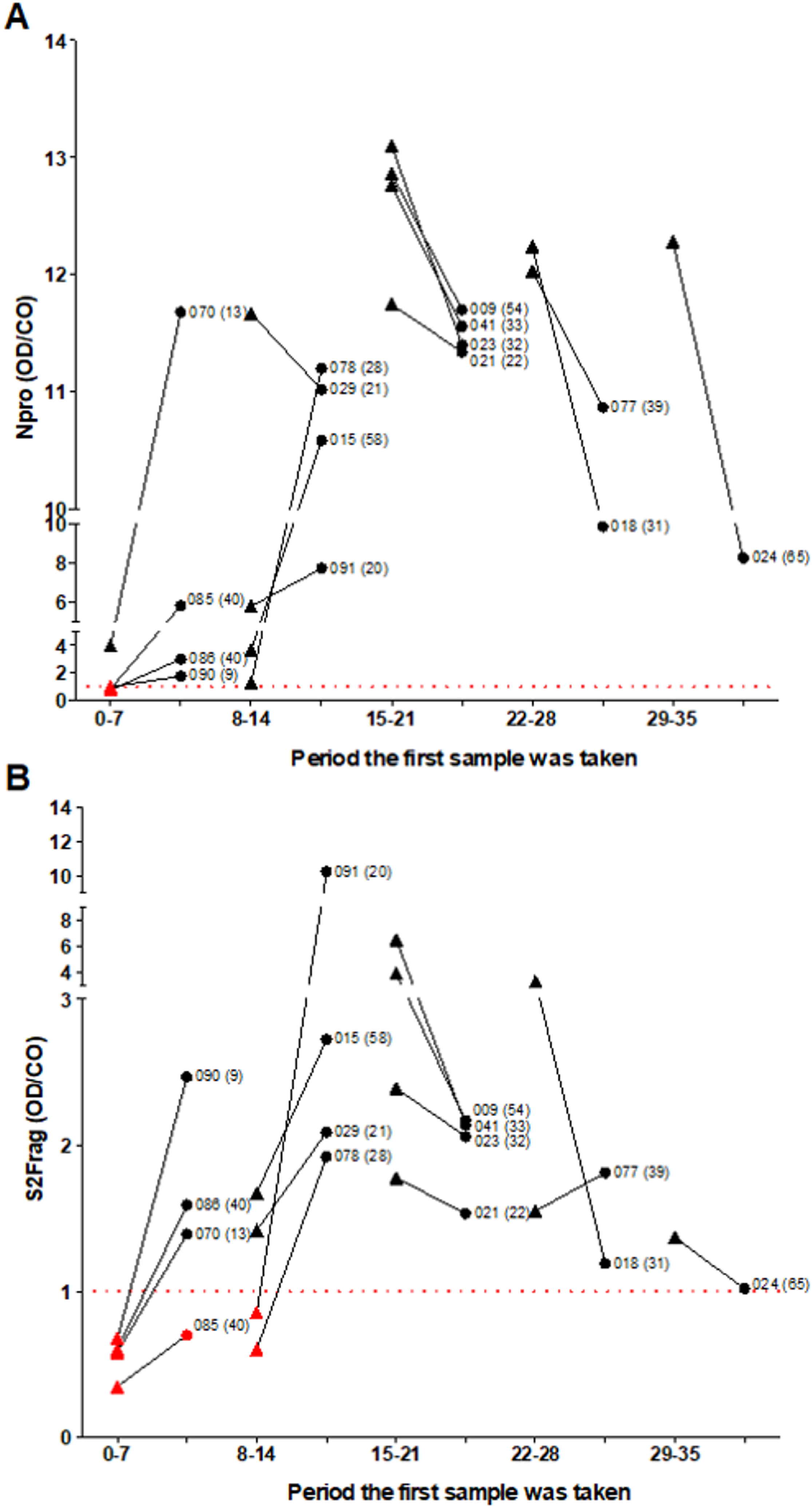
Variation of the antibody response to SARS-CoV-2 antigens in COVID-19 hospitalised patients. A, COVID-19 hospital patients plasma samples were tested to their immune response to: (A) Nucleocapsid protein (Npro) and (B) Subunit 2 of spike protein (S2Frag) in ELISA assays. The antibody response of each patient was assessed at two different time points. Samples were categorised according to the day after onset of symptoms the first plasma sample was obtained, represented in the graphic by periods. The antibody levels (OD/CO) of the two samples are compared in the graphic: Triangles represent the first sample and circles represent the second sample collected. Patient code is presented next to the antibody level of the second sample. In between parentheses the number of days after onset of symptoms that the second plasma sample was obtained. OD: Optical density at 450 nm. CO: Cut-off calculated for the specific test. The cut-off value for each antigen is indicated by the dotted line.

When considering the number of days after onset of symptoms in relation to the OD/CO values, our data show that COVID-19 patients reached their highest antibody levels to virus antigens between day 15 and 21 after onset of symptoms. Surprisingly, from day 22 the antibody responses to both Npro and S2frag begin to decline (Fig 7A and B). Since the S2Frag stimulates a weaker response, specific antibodies to this antigen dropped to levels close to the cut-off of the test within approximately seven weeks; the average S2Frag OD/CO values for the first plasma samples obtained between days 15-21 and between 28-35 were 3.64 and 1.34, respectively. These values varied less when we assessed the responses to Npro; OD/CO values varied from 12.67 to 12.2 when the same intervals were considered (Supplementary Table S1).

## DISCUSSION

Individuals infected with coronaviruses mount an immune response with protective neutralizing antibodies for a period of time [23]. Recent studies have shown that neutralizing antibodies to SARS-CoV-2 proteins can be detected in all infected individuals by day 14 after onset of symptoms [8, 24]. Both the Spro and Npro are highly immunogenic structural proteins capable of generating such an antibody response [13, 25-27]. Upon infection, the Spro is readily presented to the host as part of the invasion process. In contrast, the Npro integrates with the host cell nucleus and nucleolus and is abundantly expressed during infection, playing important roles in the transcription and replication of viral RNA and packaging of the encapsulated genome into virions [28, 29].

Since the start of the COVID-19 global pandemic, Spro and Npro have been extensively used to develop the antibody tests to diagnose past-infection by SARS-CoV-2. As antibody tests identify historic infections, they are a highly prized tool for epidemiological studies that track the spread of the virus within the community and for estimating herd immunity. However, independent and more extensive assessment of these tests has highlighted serious issues with their sensitivity that result in up to 20% false negativity [17, 18].

It has been shown that antibodies targeted against Npro appear earlier than those against Spro [30], offering an explanation as to why Npro is the antigen of choice in most commercially available tests. To understand how individuals naturally infected with SARS-CoV-2 respond to the main viral antigens, six viral proteins were recombinantly expressed: the full-length Spro and four different sub-segments, i.e. S1Frag, S2Frag, S2Prime and RBD (Fig 1), and the Npro. Through Western blot analysis, variability in the immune response to each antigen between individuals was observed (Supplementary Fig S2). At least 85% of the COVID-19 positive individuals tested in this study showed a consistent and strong antibody response to Npro. However, our data shows that 7% of the COVID-19 non-hospitalized individuals confirmed positive by qRT-PCR were misdiagnosed as negative when using either our in-house Npro-ELISA or the commercial ARCHITECT test, demonstrating that some individuals do not produce antibodies to Npro or, alternatively, had not produced these at the time of sampling.

Despite previous reports stating that the receptor-binding domain (RBD) of the Spro is highly immunogenic and the target of many neutralizing antibodies, the RBD protein produced in this study was not immunogenic (Supplementary Fig S2) [31-33]. It is worth noting that our antigens were recombinantly produced using a prokaryotic *E. coli* system, while the immunogenic recombinant RBD produced by Amanat et al. [15] was expressed in mammalian cells. This could have resulted in proteins with different antigenic properties that affect the ability of host antibodies to recognize the antigen. Nevertheless, our study agrees with Robbiani et al. [34], who observed that convalescent plasma samples from individuals who recover from COVID-19 do not contain high levels of RBD-specific neutralizing antibodies.

Conversely, the full-length Spro was consistently recognized by antibodies from individuals infected by SARS-CoV-2 [32, 35, 36]. Although the subdomain S1 protein (S1Frag), containing the RBD, is the most common fragment of the Spro used in commercial serological tests, our study found that a stronger immune response was directed against the subdomain S2 protein (S2Frag); 38 of the 42 (90.5%) individuals that were SARS-CoV-2 RT-PCR-positive elicited antibodies to the S2Frag, indicating the diagnostic value of the domain. However, based on the OD/CO values obtained, COVID-19 hospitalized and non-hospitalized positive individuals mounted stronger immune response against Npro, indicating that S2Frag is less immunogenic.

It was reported that during COVID-19 infection a decrease in the number of viral particles coincides with the appearance of neutralizing antibodies [37], although the longevity of such antibodies is debatable. Antibody titres to SARS-CoV-2 proteins were demonstrated to remain elevated for variable periods, seven days to more than 48 days, and serve to protect the individual against reinfection [8, 24]. In our study we found that infected individuals did not sustain high antibody levels to SARS-CoV-2 antigens for long periods, and even individuals that developed severe disease and required intensive care exhibited antibody declines, mainly those specific to S2Frag, after three weeks (Fig 7 A and B and Supplementary Table S1). As both anti-Spro and anti-Npro IgG antibodies have been observed to neutralize SARS-CoV-2 [38], our tests may be of use for assessing protection after infection or immunization.

By performing a dual ELISA with Npro and S2Frag we detected anti-viral antibodies in 40 out of 42 PCR-positive individuals. Follow-up Western Blot analysis of the two negative samples by ELISA, indicated that one individual had no antibodies against the viral antigens (Study code C11, supplementary Fig S4), whilst the second patient had only a weak response to S2Frag (Study code C86, supplementary Fig S4). The results obtained for C11 suggest that the patient received a false-positive qRT-PCR result, though it is important to consider that little is known about seroconversion during SARS-CoV-2 infection. While some patients may seroconvert, others might develop low antibody titres that wane within a short period of time, generating false-negative results [15]. On the other hand, the analytical sensitivity of SARS-CoV-2 qRT-PCR tests is 80% [39, 40], leaving a large potential for false negative results that we certainly observed in our study. Among the ten SARS-CoV-2 qRT-PCR negative hospitalized patients we evaluated, four tested positive for antibodies to Npro and/or S2Frag in our ELISAs (Supplementary Table S1). Our results indicate that targeting the antibody response against both Npro and S2Frag in serological diagnostic tests increases the sensitivity of detection of true positive SARS-CoV-2 infection and, therefore, represents an important strategy to improve COVID-19 diagnosis.

Our ELISAs results also revealed that ∼17% of the 42 qRT-PCR-positive individuals recognize either Npro or S2Frag antigen only; ∼7% of the individuals exclusively recognized Npro while 10% only recognised S2Frag alone (Fig 4A). These antigen-selective immune responses were confirmed using Western blot analysis (Fig 5). A similar observation was reported by Liu et al. [35], who evaluated the IgM and IgG antibody responses of 214 COVID-19 positive patients; Npro- or Spro-based ELISA resulted in positive rates of 80.4% and 82.2%, respectively, whereas together these detected 86.9% (186 patients). While these results indicate the diagnostic value of the antigens association, the differential reactivity of the serum samples with Npro and Spro was not assessed in that particular study [35].

When we analysed the antibody response of 15 patients that were hospitalized with COVID-19 we found that the Npro OD/CO values for ICU and non-ICU patients were 10.34 and 6.82 (*P* < 0.05), respectively (Supplementary Table S1), while the values for S2Frag did not vary significantly between each group (OD/CO = 1.93 and 2.22, respectively). Sun et al. [41] also found that anti-Npro IgG antibodies were significantly higher in ICU patients compared to non-ICU patients. Therefore, anti-Npro antibodies could be an indicator of disease severity, although we did not find a correlation between antibody levels and age of the patients in our study (Supplementary Fig S3).

In the present study, we compared our ELISAs results with the commercially-available immunoassay ARCHITECT (Abbott), which detects antibody response solely to Npro. The results of our in-house Npro-ELISA agreed 100% with the ARCHITECT test when we screened the 42 qRT-PCR positive sample set. However, only 6 of the 98 healthcare workers suspected of exposure to SARS-CoV-2 were deemed positive by ARCHITECT test, compared to 12 identified using our Npro-ELISA. This discrepancy rose to 14 when we employed the Npro and S2Frag dual ELISA, results which were confirmed by Western Blot analysis (Fig 5 and Supplementary Fig S4). A recent longitudinal seroprevalence study found a 95% prevalence of anti-SARS-CoV-2 antibodies in staff working in two hospitals in Ireland who had previously confirmed infection by PCR. Moreover, 16% of those with detectable antibodies reported never having experienced COVID-19 symptoms. Noteworthy, the study used primarily ARCHTECT test that was complemented with the Wantai SARS-CoV-2 AB ELISA (Fortress Diagnostics) and the Roche anti-SARS-CoV-2 immunoassay, improving the detection of positive cases and revealing that the real seroprevalence amongst the hospitals’ workers is between 2 and 5% higher than the number given by PCR diagnosis [11]. The importance of the diagnostic methods applied was further assessed by Rikhtegaran Tehrani et al. [36], which investigated 300 pre-epidemic samples and 100 PCR-confirmed COVID-19 samples using commercial tests such as EDI™ Novel Coronavirus COVID-19 ELISA, Euroimmun Anti-SARS-CoV-2 ELISA and PP® COVID-19 IgM/IgG System. This study found that their in-house Spro and Npro-based ELISAs performed with the highest sensitivity and specificity. In all, our results indicate that our in-house quantitative ELISA performs better than the non-quantitative ARCHITECT tests using a single Npro protein and can be improved by running the dual ELISA assay with S2Frag.

## CONCLUSIONS

COVID-19 serological testing in clinical settings relies on ELISA assays, which can be both qualitative and quantitative and thus a valuable tool in diagnosing past such infections [40]. However, the preference for rapid tests and the deficient performance of most commercially available SARS-CoV-2 serological tests may pose a serious risk to diagnostic efficacy [8, 17, 18]. Therefore, quantitative ELISA tests such as those developed in this study could be essential to understand the dynamic of individual antibody response to the virus and, consequently, plan appropriate measures of control during the COVID-19 pandemic.

In this study we evaluated two ELISA tests for detecting IgG antibodies to Npro and to the subdomain 2 of the Spro (S2Frag), and showed that by combining the tests we can improve the serological diagnosis of COVID-19 cases. Furthermore, we showed the applicability of the tests using plasma samples from hospitalized patients. Quantitative ELISA tests would allow us to assess antibody levels that are associated with protection or indicate a more recent or historic infection. As serological diagnostics of COVID-19 patients determine population-level surveillance and complement qRT-PCR and antigen tests, the optimization of antibody tests is critical to control the COVID-19 pandemic.

## Data Availability

The supplementary figures (Fig S1 to S4) and table (Table S1) mentioned in the manuscript are available in the Supplementary files uploaded.

## ACKNOWLEDGEMENTS

Dr David Fitzpatrick is acknowledged for expertise regarding Npro sequence identification for protein expression.

Dr Jean Dunne and Dr Niall Conlon (Department of Immunology - St James’s Hospital, James’s Street, Dublin – Ireland), Ms Fiona O’Rourke, Ms Yvonne Lynagh, Ms Martina Kelly and Dr Brendan Crowley (Department of Microbiology - St James’s Hospital, James’s Street, Dublin – Ireland) for their contributions recruiting samples, collecting data from Healthcare workers (St James’s Hospital) and analysing the samples.

## SUPPLEMENTARY FILE

Supplementary file data archive available on the Cambridge University Press - Cambridge Core website.

