## Supplementary material for "Improved diagnosis of SARS-CoV-2 by using Nucleoprotein and Spike protein fragment 2 in quantitative dual ELISA tests": Fig S1

**SUPPLEMENTARY DATA**

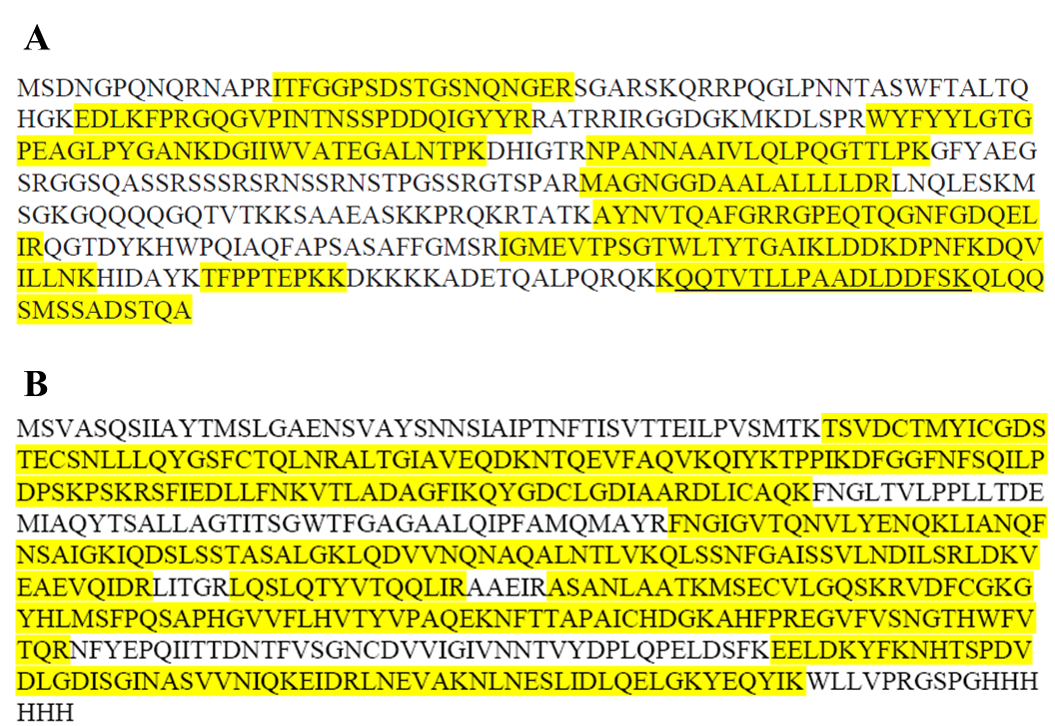

**Supplementary Figure S1.** Mass spectrometry analysis (MS/MS) to confirm the SARS-CoV-2 protein sequences recombinantly expressed. A: Nucleocapsid protein (Npro). B: Subunit 2 of Spike protein (S2Frag). The tryptic peptides that match the known sequence of amino acids for the SARS-CoV-2 identified by MS/MS are highlighted in yellow, and cover 53% and 68% of the Npro and S2Frag sequence, respectively.
