## Supplementary material for "Improved diagnosis of SARS-CoV-2 by using Nucleoprotein and Spike protein fragment 2 in quantitative dual ELISA tests": Fig S2

**SUPPLEMENTARY DATA**

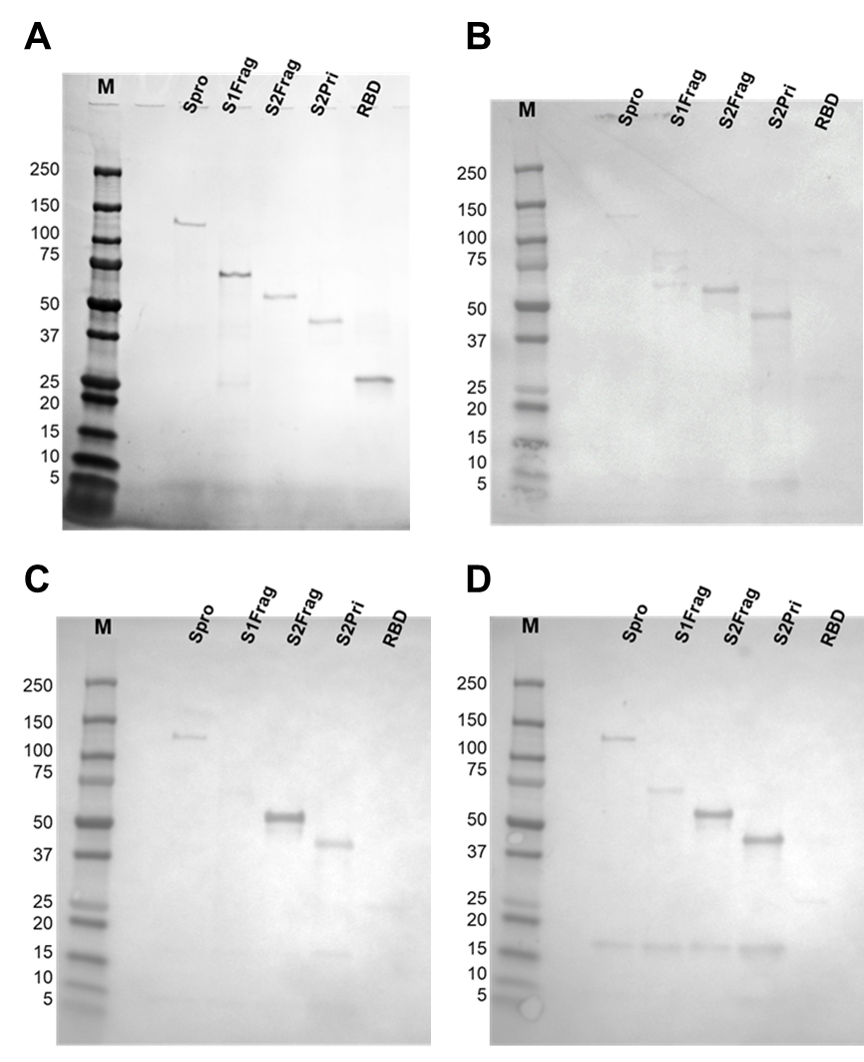

**Supplementary Figure S2. The variability of the antibody response to viral proteins of individuals positive for SARS-CoV-2.** A, Recombinant proteins resolved in a 4-12% SDS-PAGE and stained with Coomassie-blue. B – D, western-blots showing the antibody response of different SARS-CoV-2 infected individuals to different viral protein. M, molecular weight in kDa. Spike protein (Spro), Spike protein fragment 1 (S1Frag), Spike protein fragment 2 (S2Frag), S2Prime protein (S2Pri) and Receptor binding domain (RBD).
