## Supplementary material for "Improved diagnosis of SARS-CoV-2 by using Nucleoprotein and Spike protein fragment 2 in quantitative dual ELISA tests": Fig S3

**SUPPLEMENTARY DATA**

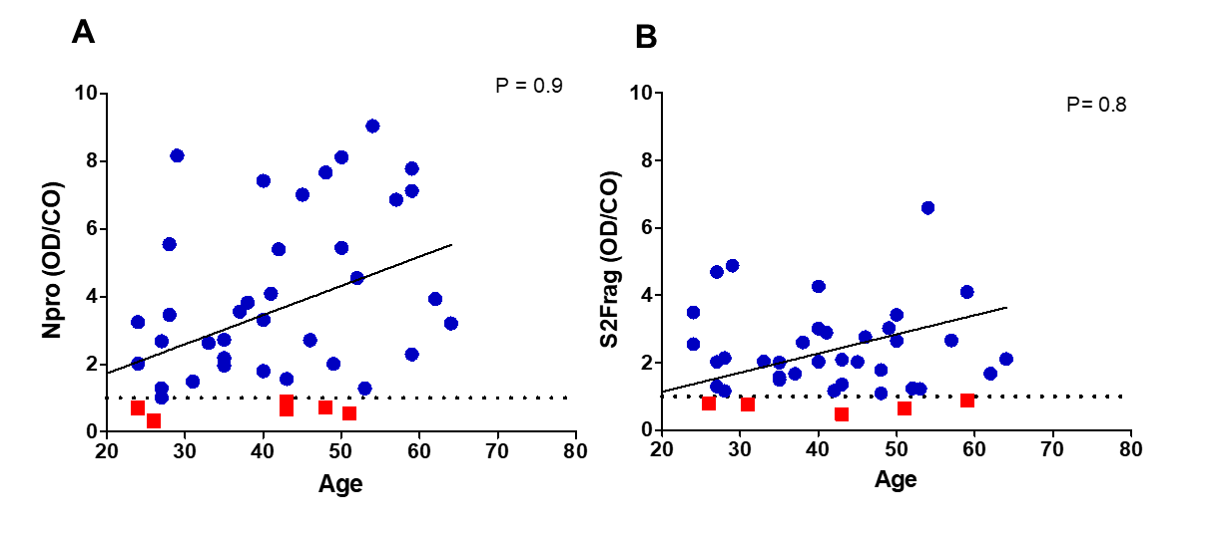
 **Supplementary Figure S3. Correlation between antibody response to SARS-CoV-2 antigens and age in 42 RT-PCR-confirmed COVID-19 individuals.** Correlation analysis (Spearman test) with 95% confidence interval. P values are shown in the figure.
