## Supplementary material for "Improved diagnosis of SARS-CoV-2 by using Nucleoprotein and Spike protein fragment 2 in quantitative dual ELISA tests": Fig S4

**SUPPLEMENTARY DATA**

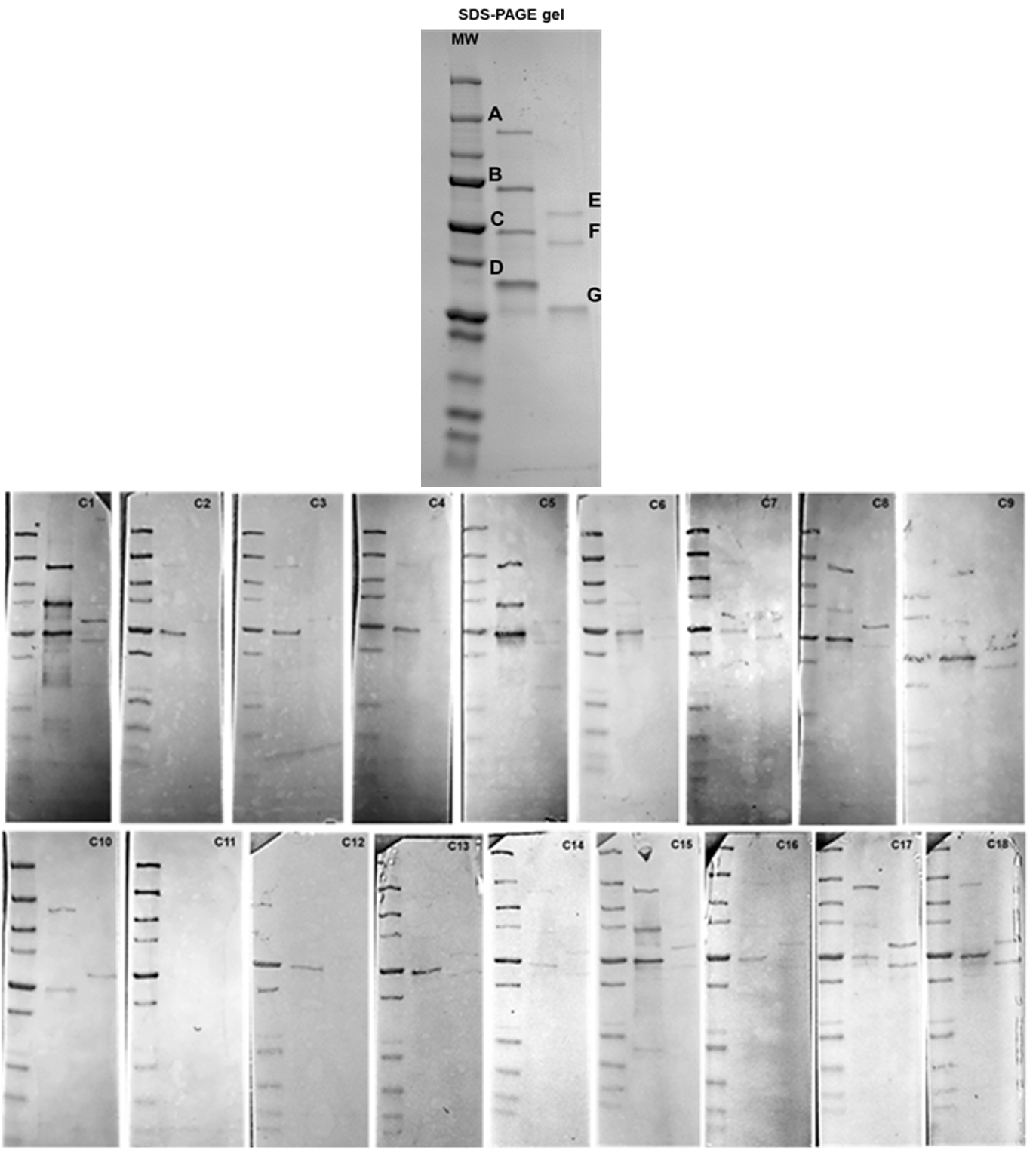

**Supplementary Figure S4A. Western blots analyses of serum samples from COVID-19 confirmed cases.** The recombinant proteins were resolved in a SDS-PAGE gel and stained with Coomassie blue as a control for the western blots. A, Spike protein (Spro); B, Spike protein fragment 1 (S1Frag); C, Nucleocapsid protein (Npro); D, 3Clike protease (Main protease) (internal control); E, Spike protein fragment 2 (S2Frag); F, S2Prime protein (S2Pri); G, Receptor binding domain (RBD). MW, Molecular weight marker in kDa. C1-C18, individual study code for serum samples used in the Western blot analyses.

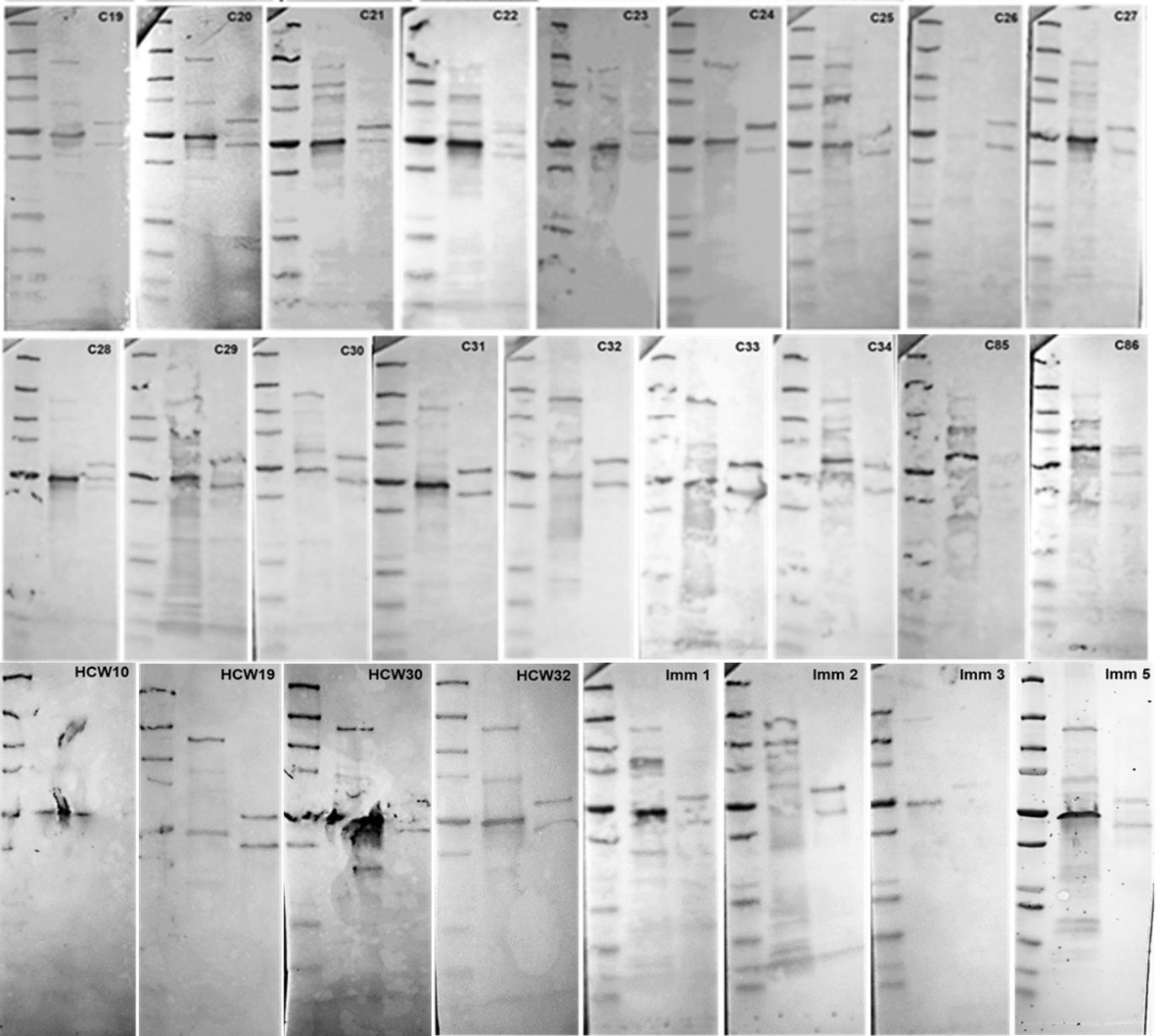

**Supplementary Figure S4B. Western blots analyses of serum samples from COVID-19 confirmed cases.** The recombinant proteins were resolved in a SDS-PAGE gel and stained with Coomassie blue as a control for the western blot, according Supplementary Fig S4A. A, Spike protein (Spro); B, Spike protein fragment 1 (S1Frag); C, Nucleocapsid protein (Npro); D, 3Clike protease (Main protease) (internal control); E, Spike protein fragment 2 (S2Frag); F, S2Prime protein (S2Pri); G, Receptor binding domain (RBD). MW, Molecular weight marker in kDa. C19-C34, C85-C86, HCW10, HCW19, HCW30, HCW32, Im1-3 and Im5, individual study code for serum samples used in the Western blot analyses.
