## Supplementary material for "Improved diagnosis of SARS-CoV-2 by using Nucleoprotein and Spike protein fragment 2 in quantitative dual ELISA tests": Table S1

**SUPPLEMENTARY DATA**

| Supplementary Table S1. Antibody levels to Npro and S2Frag in COVID-19 hospitalised patients | | | | | | | | | | |
| --- | --- | --- | --- | --- | --- | --- | --- | --- | --- | --- |
| Sample | **1^st^ sample**  **(DAO)** | **Npro ELISA** | **S2Frag ELISA** | **2^nd^ sample**  **(DAO)** | **Npro ELISA** | **S2Frag ELISA** | **RT-PCR** | **Sex** | **Age** | **ICU** |
|  |  | OD/CO | OD/CO |  | OD/CO | OD/CO |  |  |  |  |
| 009 | 17 | **12.86** | **3.89** | 54 | **11.70** | **2.17** | Positive | M | 71 | Yes |
| 015 | 11 | **3.69** | **1.67** | 58 | **10.59** | **2.73** | Positive | M | 69 | Yes* |
| 018 | 22 | **12.24** | **3.26** | 31 | **9.86** | **1.19** | Positive | M | 66 | Yes* |
| 021 | 16 | **11.75** | **1.78** | 22 | **11.34** | **1.53** | Positive | M | 65 | Yes* |
| 023 | 18 | **13.19** | **2.39** | 32 | **11.41** | **2.06** | Positive | F | 56 | Yes* |
| 024 | 33 | **12.28** | **1.37** | 65 | **8.27** | **1.02** | Positive | F | 66 | Yes* |
| 029 | 11 | **11.67** | **1.42** | 21 | **11.02** | **2.09** | Positive | M | 42 | No* |
| 041 | 20 | **12.76** | **6.52** | 33 | **11.56** | **2.14** | Positive | M | 80 | No |
| 070 | 7 | **3.97** | 0.58 | 13 | **11.68** | **1.39** | Positive | M | 53 | Yes |
| 077 | 24 | **12.03** | **1.55** | 39 | **10.87** | **1.81** | Positive | M | 62 | No |
| 078 | 9 | **1.28** | 0.60 | 28 | **11.20** | **1.93** | Positive | F | 35 | No* |
| 085 | 0 | 0.85 | 0.35 | 40 | **5.81** | 0.70 | Positive | F | 71 | No |
| 086 | 0 | 0.76 | 0.60 | 40 | **3.01** | **1.59** | Positive | M | 79 | No |
| 090 | 7 | 0.97 | 0.67 | 9 | **1.76** | **2.47** | Positive | F | 44 | No |
| 091 | 12 | **5.79** | 0.85 | 20 | **7.72** | **10.26** | Positive | F | 70 | No |
| 4-024 | - | - | - | - | 0.35 | 0.30 | Negative | F | 78 | Ni |
| 4-028 | - | - | - | - | 0.36 | 0.47 | Negative | M | Ni | Ni |
| 4-043 | - | - | - | - | 0.54 | 0.91 | Negative | M | Ni | Ni |
| 4-064 | - | - | - | - | 0.88 | 0.84 | Negative | F | Ni | Ni |
| 4-074 | - | - | - | - | 0.83 | **3.08** | Negative | F | Ni | Ni |
| 4-089 | - | - | - | - | **1.30** | 0.80 | Negative | M | Ni | Ni |
| 4-090 | - | - | - | - | **8.84** | 0.65 | Negative | F | Ni | Ni |
| 4-118 | - | - | - | - | 0.55 | 0.41 | Negative | F | Ni | Ni |
| 4-120 | - | - | - | - | 0.52 | 0.74 | Negative | F | Ni | Ni |
| 4-231 | - | - | - | - | **1.32** | 0.93 | Negative | F | 89 | Ni |
| DAO: number of days after the onset of symptoms. OD: Optical density at 450 nm. CO: Cut-off of the specific ELISA. OD/CO >1: Positive for ELISA; OD/CO <1: Negative for ELISA. ICU: Intensive care unit. (*): Patients that required invasive ventilation. Ni. Not informed. | | | | | | | | | | |
